# Exploring the Utility of a Functional Magnetic Resonance Imaging (fMRI) Cannabis Cue-Reactivity Paradigm in Treatment Seeking Adults with Cannabis Use Disorder

**DOI:** 10.1101/2023.11.14.23298485

**Authors:** Gregory L. Sahlem, Logan T. Dowdle, Nathaniel L. Baker, Brian J. Sherman, Kevin M. Gray, Aimee L. McRae-Clark, Brett Froeliger, Lindsay M. Squeglia

## Abstract

**Introduction:** Functional magnetic resonance imaging (fMRI) studies examining cue-reactivity in cannabis use disorder (CUD) to date have either involved non-treatment seeking participants or been small. We addressed this gap by administering an fMRI cue-reactivity task to CUD participants entering two separate clinical trials.

**Methods:** Treatment-seeking participants with moderate or severe CUD had behavioral craving measured at baseline via the Marijuana Craving Questionnaire (MCQ-SF). They additionally completed a visual cannabis cue-reactivity paradigm during fMRI following 24-hours of abstinence from cannabis. During fMRI, the Blood Oxygen Level Dependent (BOLD) signal was acquired while participants viewed cannabis-images or matched-neutral-images. BOLD responses were correlated with the MCQ-SF using a General Linear Model.

**Results:** N=65 participants (32% female; mean age 30.4±9.9SD) averaged 46.3±15.5SD on the MCQ-SF. When contrasting cannabis-images vs. matched-neutral-images, participants showed greater BOLD response in bilateral ventromedial prefrontal, dorsolateral prefrontal, anterior cingulate, and visual cortices, as well as the striatum. Similarly, there was stronger task-based functional-connectivity (tbFC) between the medial prefrontal cortex and both the amygdala and the visual cortex. There were no significant differences in either activation or tbFC between studies or between sexes. Craving negatively correlated with BOLD response in the left ventral striatum (R^2^=-0.25; *p*=0.01).

**Conclusions:** We found that, among two separate treatment-seeking CUD groups, cannabis cue-reactivity was evidenced by greater activation and tbFC in regions related to executive function and reward processing, and craving was negatively associated with cue-reactivity in the ventral striatum. Future directions include examining if pharmacological, neuromodulatory, or psychosocial interventions can alter corticostriatal cue-reactivity.

## Introduction

Approximately 2% of the United States population meets criteria for Cannabis Use Disorder (CUD) and the prevalence may be increasing^1,2^. Concordantly there is a high demand for treatment; however, despite much effort, current treatment options have limited efficacy^3^. To date the most established and efficacious treatments are based on behavioral approaches^4^, with several medications also showing promise^5–8^, but no medication yet proven to be definitively efficacious. In a similar fashion to clinical trials in other substance and psychiatric disorders, there is little insight into which patients will or will not respond to any given treatment, and there are little data examining the neural mechanism of action of any given treatment for CUD.

Given the phenotypic heterogeneity of those with CUD and the low rates of abstinence following quit attempts, there has been increasing interest in examining CUD (along with other substance use disorders and psychiatric illnesses) from a neural basis, whereby each illness is characterized by dimensional behavioral dysfunction, which is then associated with the underlying neural systems that govern them. The current framework for this approach was developed and described in the National Institutes of Mental Health’s Research Domain Criteria, or RDoC initiative^9^. A similar framework for a neural approach to addictive disorders was developed in parallel and remains the prevailing heuristic, whereby distinct neural systems subserve distinct behavioral dysfunction, which converge to result in the addictive end phenotype^10,11^. To systematically study substance use disorders in the context of their neural etiology, further work is needed to establish and validate neural targets that directly relate to relevant dysfunctional behaviors.

Drug craving and the related response to drug cues (cue-reactivity) are the behavioral constructs in addictions that have been studied most thoroughly. Craving has been a frequent proximate target in clinical trials, and has clear clinical relevance in opioid^12,13^, alcohol^14,15^, nicotine^16^ and stimulant use disorders^17,18^. Neuroimaging studies (predominantly using functional magnetic resonance imaging, fMRI), exploring the neural substrates of cue-reactivity have consistently found activation of incentive salience related structures— including the ventromedial cortices; the anterior and subgenual portions of the cingulate gyrus; and the striatum—in response to drug cues, relative to neutral cues^11^. Several studies have related drug cue-reactivity in these regions to clinically relevant outcomes^19^, and a number of studies have demonstrated that effective pharmacologic treatments both modulate drug cue-reactivity within these nodes (demonstrate target engagement) and group-level target engagement is associated with better clinical outcomes including among those with opioid^20,21^, alcohol^22–24^, and nicotine use disorders^25,26^. However, relatively little is known about whether the same relationship exists between engaging neural activation during cue-reactivity and clinical outcomes in CUD, and only a single trial has prospectively linked fMRI cue-reactivity to clinical outcomes in CUD^27^.

There is an extensive literature suggesting that both adults and adolescents who heavily use cannabis display behavioral cannabis cue-reactivity (cue-induced-craving)^28^. A series of neuroimaging studies have extended these behavioral findings, and have demonstrated that cannabis users display the characteristic increase in activation in incentive salience related structures in response to cannabis cues^29–37^. When taken together, these experiments suggest that addiction severity is related to the degree of fMRI activation^27,30,38,39^, neural activation correlates with clinically relevant behavioral data^27,34,38,40^, and genetics may play an important role in cue-reactivity^41^. Though relatively few studies have examined task-based network connectivity among individuals with a CUD, both^36,39^ found increased connectivity between striatal and prefrontal areas when participants viewed cues. Though these seminal investigations have provided important insights into the neural basis of cue-reactivity in heavy cannabis users, only two of the above studies recruited rigorously screened treatment seeking participants^34,40^, and the sample-size for both of those studies combined is 32, leaving this group minimally studied, and uncertainty whether neural cue-reactivity can be used as an assessment in clinical trials.

In order to better characterize the neuropathophysiology of SUDs and CUD specifically, there is a need for validated neural paradigms, that: reliably and specifically activate neural regions underlying problematic use; have clinical relevance; have good test-retest reliability; and are responsive to treatments that alter behavior. In order to take the first step in this endeavor, our group took baseline imaging data (using an fMRI task that was previously validated in non-treatment-seeking adolescents^31^) from two independently run treatment trials that each recruited and rigorously screened participants with CUD. We sought to determine if the task reliably induced the expected activation-patterns and task-based functional connectivity patterns in each study separately to determine if it is generalizable across studies (hypothesizing that it would). We then combined the data to increase our power to explore associations between task activation and behavior to determine if there were clinically-relevant relationships, such as neural activation correlations with craving, marijuana related problems, and amount of cannabis use (hypothesizing that there would be). Given this was a secondary analysis that was not pre-planned, we took an exploratory whole-brain approach with the hopes of using the findings for future investigations.

## Methods

### Overview and participant evaluations

This cross-sectional study leveraged the baseline evaluations of two clinical trials attempting to treat individuals with moderate or severe CUD who were interested in reducing their cannabis use. The two trials investigated the potential therapeutic effects of varenicline^8^ (NCT02892110), and repetitive Transcranial Magnetic Stimulation^42^ (NCT03144232) respectively. All research related activities were approved by the Medical University of South Carolina’s Institutional Review Board (IRB) and were conducted in accordance with the declaration of Helsinki. Participants in both trials were recruited via media and print advertisements from the greater Charleston, SC, area, underwent a brief phone screen assessing eligibility, and if eligible for the study, were invited for an in-person screening and enrollment visit. After reviewing and signing informed consent, participants underwent a similar screening procedure in both trials which included evaluation with a brief medical examination, the Mini International Neuropsychiatric Interview (MINI^43^), the Time-Line Follow-Back (TLFB^44^), the Marijuana Problem Scale (MPS^45^), urine drug testing (UDT; Alere Toxicology, testing for amphetamines, benzodiazepines, cannabis, cocaine, and opiates), and creatinine-corrected urine cannabinoid testing (mg/ml) derived by dividing the urine cannabinoid levels (minimum detection cut-off value of 30.00 ng/ml, Abbott AXSYM®) from urine creatinine (mg/ml).

Overlapping inclusion criteria for both studies included: a) age 18-55 (or 18-60 for rTMS); b) currently meeting DSM-5 criteria for ≥moderate CUD and cannabis use ≥3 days per week in the last 30 days (≥5-days/week for rTMS); c) interest in quitting or decreasing cannabis use; and d) sufficient intellectual level and command of the English language to provide consent and complete assessments. Additionally, participants in the varenicline study had to have a body mass index between 18 and 35kg/m^2^ and a weight greater than 50kg for pharmacokinetic reasons. Overlapping exclusion criteria included: a) being pregnant or breastfeeding; b) currently meeting DSM-5 criteria for ≥moderate non-cannabis/tobacco substance use disorder; c) current unstable psychiatric, neurologic, or general medical condition; d) lifetime history of bipolar or psychotic disorder; e) active suicidal ideation, or a suicide attempt within the past 90 days (120 days for varenicline); f) unstable dosing for central nervous system medications (no central nervous system active medications for rTMS); or g) contraindications for MRI such as claustrophobia, or implanted metal. Additionally, participants were excluded from the rTMS study if they had a history of seizure and were excluded from the varenicline study if they had taken an investigational agent in the last 30 days or were enrolled in another clinical trial within 60 days.

Participants meeting the above inclusion/exclusion criteria were invited back for a scanning visit where they were asked to abstain from the use of cannabis or other drugs for at least 24 hours (verified by self-report and a Confirm Biosciences saliva test for amphetamines, benzodiazepines, cocaine, cannabis, and opiates). Behavioral craving was assessed using the short form of the Marijuana Craving Questionnaire (MCQ-SF)^46^, prior to scanning in the case of the varenicline study, and approximately 20-minutes after scanning in the case of the rTMS study. Of note the MCQ-SF was collected only once (as opposed to before and after the fMRI). Urine drug testing was also performed, and urine cannabinoids were quantified using a standard assay and creatinine correction (see above).

### Imaging Procedures

For this investigation, MRI was conducted using a 32-channel head coil with a 12-m gradient-echo, echoplanar imaging (EPI) sequence on a 3-Tesla Prisma^fit^ MRI scanner (Siemens, Erlangen, Germany). We first collected a Scout image to align subsequent image acquisitions. Next, we collected an anatomical image (MPRAGE, 1mm^3^ TR 2300ms, TE 2.26ms, TI 900ms) for functional alignment and transformation to standard Montreal Neurological Institute (MNI) space. Then we collected functional data while participants completed a visual cannabis cue task (51 slices, MB factor 3, TR 1200ms, TE 30ms, FA 65, 2.8x2.8x2.8mm, 601 volumes). Forward and reverse spin echo sequences were also collected for distortion correction.

We employed a previously developed block-design cannabis cue task^31^. During the task, a series of high-resolution cannabis-images, matched-neutral-images, blurred-images, or a fixation crosshair were presented on a projection screen visible to the participants via a mirror attached to the head coil. Images were presented during six 120-second epochs, each consisting of four repetitions of 24-second blocks of an image type, followed by a 6-second rating period. Each image block consisted of 12-pictures presented for two seconds each. During each rating period participants rated their current urge to use cannabis on a 5-point Likert scale (1-None to 5-Extreme), using a button-push hand pad. The images consisted of 36 cannabis-images and 36 matched-neutral-images of non-food objects or plants (matched on color, hue, and visual complexity). There were subsequently a total of 36 matched pairs of cannabis and neutral images. For the cannabis-images, there were two-blocks of ‘passive’ cannabis-images which included paraphernalia or a cannabis plant, and one-block of ‘active’ cannabis-images depicting individuals smoking or handling cannabis or paraphernalia. There were coinciding blocks of ‘active’ and ‘passive’ matched-neutral-images. Blurred images and fixation trials were used as contrasts to evaluate attention and non-cannabis specific effects, and the six-second rating period allowed for a normalization of the hemodynamic response in-between blocks.

### Imaging pre-processing and task activation modelling procedures

We performed fMRI preprocessing using Analysis of Functional Images (AFNI) version 6.33^47^. Anatomical data were fed through AFNI’s @SSwarper to calculate non-linear transformations into MNI space. We used afni_proc.py to perform standard preprocessing, which included despiking, slice timing adjustment, motion correction, distortion correction using images with reverse phase encoding, anatomical alignment, and MNI normalization. All spatial transformations were concatenated and applied in a single step in order to reduce interpolation errors. Next, the MNI-space functional data were passed through ICA-AROMA^48^ using the ‘non-aggressive’ setting. Output data were then entered into a second afni_proc.py function call, that performed blurring (6mm FWHM Gaussian kernel) and scaled the data such that each voxel had a mean of 100.

To determine the activation associated with each type of image, we used a general linear model (GLM) approach using AFNI’s 3dREMLfit, which accounts for autocorrelations in the fMRI time-series^49^. Task event onsets were modeled in seconds and convolved with an estimate of the hemodynamic response (double gamma ‘SPMG1’) function with the appropriate duration (24-seconds for images, 6-seconds for ratings). Motion parameters and their derivatives were also included in the model, as high pass filtering with polynomial detrending up to order 5.

Group statistics from the beta estimates and t-statistics produced at the first level were calculated using a mixed modeling approach in 3dMEMA^50^. The smoothness of the data was calculated from the residual time-series using the more accurate Auto-Correlation Function (ACF) metric^51^. These values were averaged across all participants. We then used Monte Carlo estimation in order to determine cluster thresholds corresponding to a p_FWE_<0.05 for a voxel level threshold of p<0.001, bi-sided^52^. Our primary contrast of interest was that of cannabis-images vs. matched-neutral-images. In addition, we examined activity in response to cannabis-images/matched-neutral images vs. blur-images and fixation cross. We performed each of the contrasts above in each study individually, and then contrasted the two studies. We additionally performed each contrast in male and female participants and contrasted the two sexes. Finally, we also compared activation during ‘active’ cannabis-images to activation during ‘passive’ cannabis-images (‘active’ cannabis-images > ‘active’ matched-neutral-images) > (‘passive’ cannabis-images >passive neutral images) to determine the effects of these stimulus qualities on BOLD responses.

After the basic GLM was completed, we took a data driven approach and extracted the beta values (corresponding to % signal change) from the maximum activation point of multiple whole brain points of greater activation that were consistent with other cue reactivity trials. These included the clusters identified as the left and right ventral striatum, the ventromedial prefrontal cortex(vmPFC) / anterior cingulate cortex (ACC), the left and right visual cortex, and the right dorsolateral prefrontal cortex. These specific clusters were chosen based on their consistency with other reports^53^.

### Task-Based Functional Connectivity analysis

We used the CONN toolbox to determine the changes in functional connectivity between when participants were viewing cannabis-images to when they were viewing matched-neutral-images. First, we used the same denoised data that were entered in the GLM above, applied smoothing in CONN, used default filtering settings, and regressed motion parameters and their derivatives^48,54^. The data was band passed between 0.008 and 0.09 Hz. Time-series were extracted from the unsmoothed data using the following full MNI ROI set provided with CONN18b, which provides 132 ROIs across cortical and subcortical regions. We excluded the cerebellar ROIs (n = 27), leaving 105 ROIs in the analysis. For a full list of ROIs used, see Supplemental Table-3.

Functional connectivity was calculated using bivariate regression for each subject, with hemodynamic response function weighting and then combined in a group level analysis. We contrasted the connectivity during cannabis-images with matched-neutral-images and corrected for multiple comparisons using the strict *p*_FDR_<0.05 analysis level correction, which accounts for both the number of target and source ROIs (105). In a similar fashion to the above extracted beta values, we extracted the correlation coefficients (Rz-scores) from connections between the following data-driven regions: vmPFC to amygdala; vmPFC to occipital; vmPFC to left-lateral-occipital; vmPFC to right-lateral-occipital; and left-parietal to occipital.

### Behavioral data analysis procedures

We approached the behavioral data by first comparing demographic descriptors (age, sex, race/ethnicity, and educational achievement); duration of cannabis use (age of onset of any use); illness severity (DSM-5 Criteria and MPS); cannabis craving (MCQ-SF, and hand-pad urge rating); and cannabis use (TLFB and urine creatinine corrected cannabinoids) between studies utilizing the Wilcoxon Rank Sums test for continuous variables and Chi-Square (or Fisher’s exact when appropriate) tests for categorical variables. We then calculated Spearman rank correlation coefficients between craving (MCQ-SF total score), CUD related problems (the MPS), cannabis use (number of cannabis use sessions over the 7-days prior to scanning), the extracted activation %signal-changes in the six data-driven clusters above, and the extracted connectivity Rz-scores from the five data-driven connectivity pairs above. We further explored significant associations between cluster activation or Rz-scores, and behavioral data, using a general linear model. Continuous and normally distributed outcomes (MCQ-SF, MPS) were modeled assuming a Gaussian distribution. The number of cannabis use sessions in the prior 7-days was modeled using a negative binomial regression. In all models, the primary predictors were the activation region of interest, as well as study demographic variables (age, sex, race/ethnicity, education, marital status, and tobacco smoker status), and the dependent variable of interest was the target behavioral construct. We used backward selection to develop parsimonious models. Residual normality was assessed in Gaussian models. As there were two distinct studies included in the analysis, a fixed study effect was included in all regression models. We did not correct for multiple comparisons in the behavioral data in this preliminary investigation. Statistical significance, when reported, was based on two-tailed tests with an alpha of 0.05. All statistical analyses were run using SAS University Edition (Cary, NC, USA).

## Results

### Demographic and descriptive data (Table-1 and Supplemental Table-1)

**Table-1:** Sample Descriptive Statistics:

|  | Total Sample | rTMS | Varenicline | Difference |
| --- | --- | --- | --- | --- |
| **Simple Descriptive Variables:** |  |  |  |  |
| Age | 30.4 ± 9.9 | 31.3±11.2 | 29.8±8.8 | *p*=0.90 |
| Sex | 44 Men<br>21 Women | 18 Men<br>10 Women | 26 Men<br>11 Women | *p*=0.61 |
| Race and Ethnicity: AA = African American; C = Caucasian; and O = Other | 20 AA; 42 C; 3 O | 7 AA; 18 C; 3 O | 13 AA; 22 C; 2 O | *p*=0.24 |
| Marital Status | 9 Married<br>55 Non-Married | 2 Married<br>26 Non-Married | 7 Married<br>29 Non-Married | *p*=0.28 |
| Education level | 41 < 4-year<br>24 ≥ 4-year | 18 < 4-year<br>10 ≥ 4-year | 23 < 4-year<br>14 ≥ 4-year | *p*=0.44 |
| Smoked cigarettes on at least 14 of the previous 28 days | 20 Smokers<br>45 Non-Smokers | 9 Smoker<br>19 Non-Smoker | 11 Smoker<br>26 Non-Smoker | *p*=0.83 |
| Age of First Cannabis Use: | 15.5 ± 3.9 | 15.0 ± 2.4 | 16.0 ± 4.7 | *p*=0.46 |
| **Diagnostic and Statistical Manual Criteria (DSM-5) and Marijuana Problem Scale Values** |  |  |  |  |
| # of DSM-5 Cannabis Use Disorder Criteria met | 7.4±1.7 | 8.0±1.6 | 6.9±1.7 | ***p*=0.010** |
| DSM-5 Cannabis Use Disorder Categorical | 56 Severe<br>9 Moderate | 26 Severe<br>2 Moderate | 30 Severe<br>7 Moderate | *p*=0.28 |
| Marijuana Problem Scale score (MPS) | 9.2±5.8 | 7.8±5.4 | 10.2±6.0 | *p*=0.13 |
| **Short Form of the Marijuana Craving Questionnaire (MCQ) and fMRI hand-pad craving values:** |  |  |  |  |
| MCQ-SF-Total | 46.3±15.5 | 45.4±16.3 | 47.0±15.2 | *p*=0.64 |
| MCQ-SF Compulsivity | 9.1±4.5 | 9.1±5.3 | 9.1±4.0 | *p*=0.68 |
| MCQ-SF Emotionality | 10.8±5.4 | 9.9±5.5 | 11.4±5.2 | *p*=0.25 |
| MCQ-SF Expectancy | 12.6±4.5 | 11.8±4.6 | 13.2±4.5 | *p*=0.31 |
| MCQ-SF Purposefulness | 13.8±4.9 | 14.5±5.2 | 13.3±4.7 | *p*=0.32 |
| fMRI Handpad (cannabis – neutral) | 1.2±1.0 | 1.4±1.0 | 1.1±0.9 | *p*=0.17 |
| **Cannabis Use variables:** |  |  |  |  |
| Urine cannabinoids Level (ng/ml) | 337.4±471.5 | 346.4±557.3 | 330.3±400.6 | *p*=0.79 |
| Creatinine (mg/ml) | 139.7±86.4 | 140.2±71.3 | 139.3±98.1 | *p*=0.76 |
| Creatinine corrected cannabinoids level (ng/mg) | 3.5±4.18 | 3.1±4.4 | 3.8±4.0 | *p*=0.38 |
| # days using cannabis / last 7 | 5.1±2.3 | 4.9±2.4 | 5.2±2.3 | *p*=0.56 |
| # days using cannabis / last 28 | 24.1±5.2 | 23.3±6.2 | 24.6±4.4 | *p*=0.52 |
| # cannabis use sessions / last 7 | 15.6±16.4 | 17.6±19.8 | 14.1±13.4 | *p*=0.79 |
| # cannabis use sessions / last 28 | 86.5±67.2 | 89.9±84.1 | 84.3±54.6 | *p*=0.57 |

All values reported ± Standard Deviation (SD).

The final sample included a total of 65 participants (twenty-eight in the rTMS study and thirty-seven in the varenicline study). The average age of included participants was 30.4±9.9SD and consisted of 32.3% women. There were no significant differences in age, sex, or other demographic variables between the two studies (see Table-1 for additional details). There was, however, a significant difference between the rTMS and varenicline samples regarding DSM-5 criteria (participants in the rTMS group met on average 8.0±1.6SD DSM-5 criteria, while those in the varenicline study met on average 6.9±1.7SD criteria; p=0.01).

### Task activation modelling

Our primary contrast of interest was cannabis-images vs. matched-neutral-images. There was greater neural activation during cannabis-images in the bilateral ventromedial prefrontal cortices, anterior cingulate cortices, striatum, visual cortices, and dorsolateral prefrontal cortices relative to during matched-neutral-images. During matched-neutral-images, there was greater activation in bilateral cuneus, right Rolandic operculum, superior temporal gyrus, and right middle cingulate cortex relative to cannabis-images (Figure-1, Figure-2, Supplemental Table-1, and Supplemental Figure-1). There were no differences between study or sex contrasts in the whole-brain analysis. However, the extracted %signal change between the cannabis-cues vs. matched-neutral-cues contrast in both the left and right ventral striatum differed between the two studies. Right ventral striatum %-signal change was 0.067±0.087SD in the rTMS group and 0.021±0.015SD in the varenicline group; p=0.05. Left ventral striatum %-signal change was 0.058±0.078SD in the rTMS group, and 0.017±0.080SD in the varenicline group; p=0.05).

**Figure-1:**
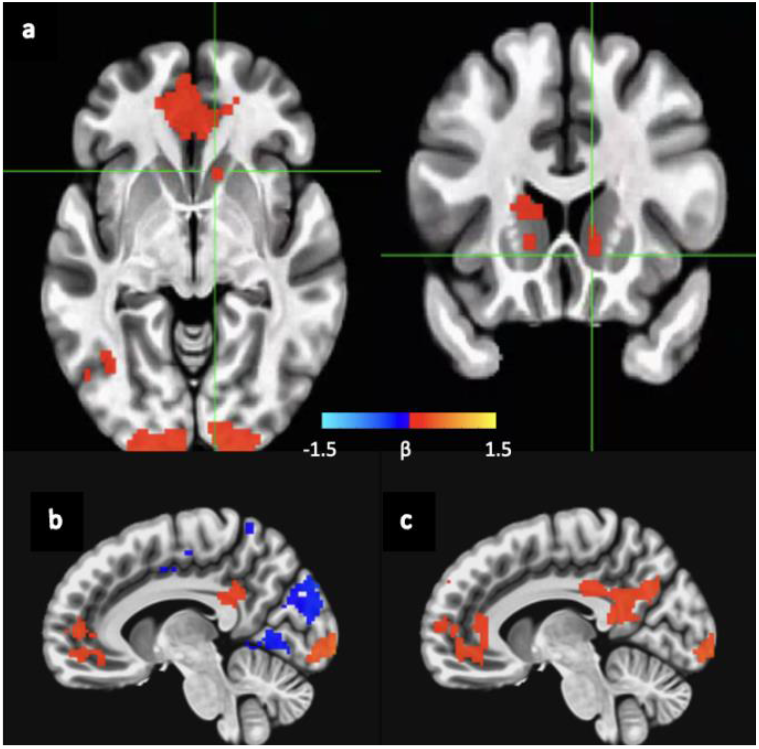
Subtraction maps contrasting cannabis-images and matched-neutral-images in: a) the combined sample (N=65); b) the varenicline sample (N=37), and; c) the rTMS sample (N=28). All comparisons used a voxel threshold of p<0.005, a comparison of p<0.001; a cluster significance of p<0.05, and; were family wise error (FWE) corrected. Critical T-statistic for the subtraction map is 3.4491 for p <0.001, two-sided. Significant clusters had to exceed 31 voxels. There were no significant differences when contrasting images b) and c) finding that the subtraction maps of the two samples did not differ significantly.

**Figure-2:**
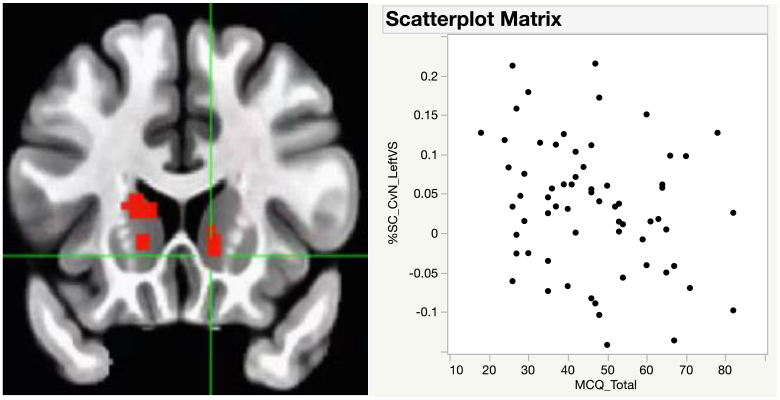
Correlation between the left ventral striatal cluster (derived from the whole brain analysis) and Marijuana Craving Questionnaire Short Form (MCQ-SF) score for the combined sample (R^2^=0.25; *p*=0.01).

When contrasting ‘active’ vs. ‘passive’ cannabis-images with ‘active’ vs ‘passive’ matched-neutral-images there was greater activation across five clusters including the bilateral inferior occipital cortex, fusiform gyrus, lateral occipital cortex, middle occipital gyrus and the left inferior parietal lobule. Lower activation was found in one cluster—spanning the left calcarine and lingual gyrus (Supplemental Figure-2 and Supplemental Table-2).

### Task-based functional connectivity modelling

During our primary comparison of interest (cannabis-images vs. matched-neutral-images) there was greater connectivity between the vmPFC to amygdala, the vmPFC to occipital, the vmPFC to left-lateral-occipital, the vmPFC to right-lateral-occipital, and the left-parietal to occipital regions (Figure-3). All connectivity changes were verified as positive (greater connectivity) when viewing cannabis-images relative to matched-neutral-images.

**Figure-3:**
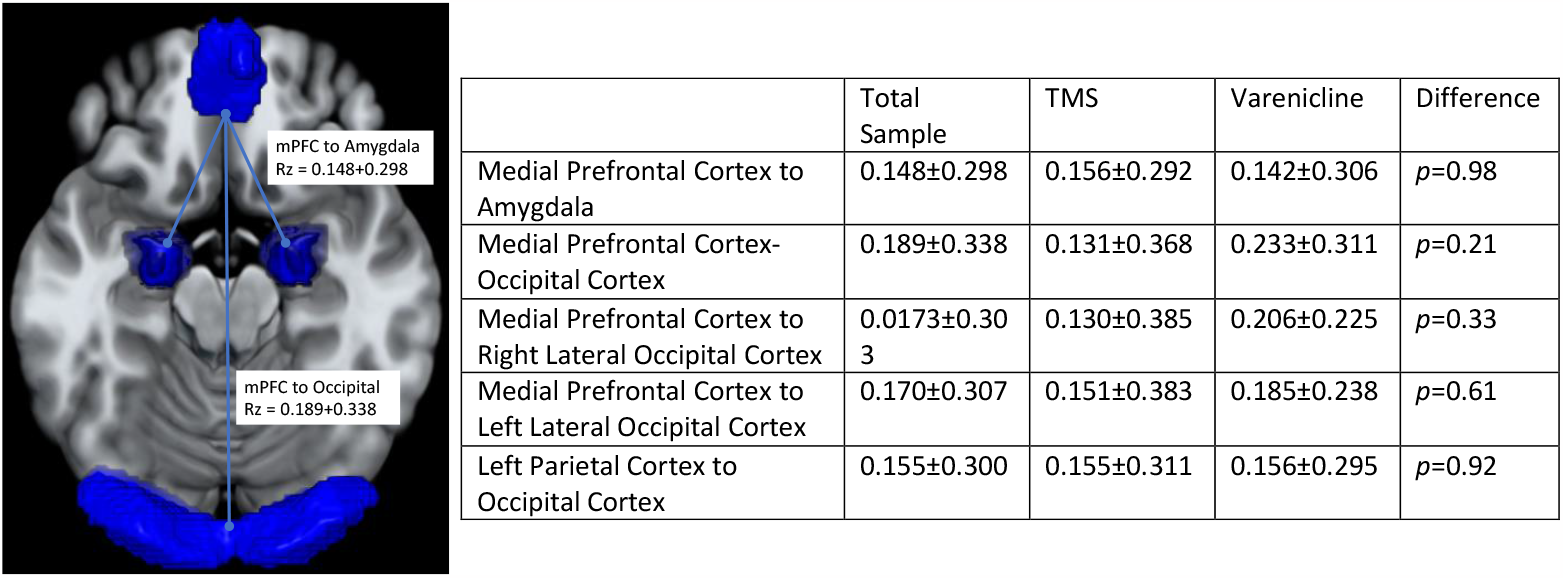
Task Based Functional Connectivity contrasting connectivity during cannabis-images (both active and passive) relative to matched-neutral-images for the combined sample. All values represent Rz values between the denoted regions of interest reported ± Standard Deviations (SD). See Supplemental Table-3 for the Conn ROI atlas that was used.

### Covariate analysis between imaging and behavioral findings

When not adjusted for covariates, the MCQ-SF total score was negatively correlated with activation in the left ventral striatum (rho=-0.26; p<0.04) and right visual cortex (rho=-0.29; p<0.02), and; the total number of cannabis use sessions over the 7-days prior to scanning was negatively correlated with percent signal change in the right DLPFC cluster (rho=-0.27; *p*=0.03). When controlling for sex, study, and tobacco smoker status, left striatal activation was still significantly associated with the MCQ-total score (R^2^=0.25; *p*=0.01). Percent signal change in the right visual cortex was no longer significantly associated with the MCQ-SF total score in adjusted models. When controlling for study and smoker status, percent signal change in the R-DLPFC was no longer significantly associated with the number of self-reported use sessions over the past 7-days. There were no significant correlations between any of the examined behavioral constructs and task-based functional connectivity.

## Discussion

In this trial, we present preliminary cue-reactivity data from two cohorts of rigorously screened participants with CUD who were entering treatment trials. Broadly, we found task-activation in structures consistent with the existent cue-reactivity literature across substance use disorders. Importantly we found consistent task-activation in both cohorts independently and did not find any significant activation differences between studies. More specifically we found that during presentation of visual cannabis-images contrasted with matched-neutral-images there was greater bilateral BOLD activation in the ventromedial prefrontal cortices, dorsolateral prefrontal cortices, anterior cingulate cortices, striatum, and visual cortices. BOLD activation in the left striatum negatively correlated with spontaneous craving as measured by the MCQ-SF collected on the same day. We further found that when comparing active cannabis to passive cannabis cues, and controlling for active vs passive neutral stimuli, as expected, we found greater activation in multiple regions, including secondary visual areas. Finally, we found that there was greater connectivity between the medial prefrontal cortex and both the amygdala and visual cortices when participants were viewing cannabis relative to matched neutral images. We did not find any differences when comparing male and female participants. We explore each of these findings in the context of the existent literature in CUD, and other substance use literature where appropriate.

The main finding of this investigation is that in two independently run CUD treatment trials, there was robust neural activation in incentive-salience and visual cortices (using a data driven whole brain approach), and that activation in the left ventral-striatum negatively correlated with baseline behavioral cannabis craving even when adjusting for study, sex, and smoker status. Each of the independent samples exceeded the sample size of the two other studies which included treatment seeking participants with CUD and found similar activation patterns relative to the existent literature. Our findings also replicate the common finding that there is more activation in the visual cortex during cue presentation as compared to neutral presentation^55^. Our findings are important for two reasons. First, the findings observed in studies which included non-treatment seeking participants are strengthened given our similar findings in treatment seeking participants with CUD. Second, these findings provide a starting point for future interventional trials for both predictive and target engagement goals. Ideally future investigations will have repeated fMRI observations and can explore test-retest consistency, and then whether interventions with behavioral effects are able to engage circuitry in a meaningful fashion (similar to the approach of Karoly and colleagues in non-treatment seeking adolescents^56^). To date there are no published trials with treatment-seeking participants with CUD that explore either of these questions, however studies in other substance use disorders^24^ suggest visual cue-reactivity may be useful. The finding that left-ventral striatum activation negatively correlated with behavioral craving (measured via the MCQ-SF) supports the findings of^38^ and contrasts the findings of^40^. Of note there were methodologic differences between studies, which include a whole brain approach in the present study and an ROI approach in the two other studies, and; treatment-seeking participants with CUD with 24-hours of abstinence in the present trial, while the other treatment-seeking trial^40^ scanned participants while they were using cannabis ad libitum and found a positive correlation, and the non-treatment seeking trial^38^ required at least 24-hours of abstinence, and found a negative correlation with the cohort of heavy cannabis users (likely to meet CUD criteria). The period of abstinence prior to scanning may subsequently be a critical component of the relationship between striatal activation and craving.

When comparing connectivity during cannabis cues and matched neutral images, we found higher connectivity between the vmPFC and both the visual cortex and the amygdala. Our findings contrast somewhat with the two other trials examining task-based functional connectivity in cannabis users^36,39^, which found greater connectivity between the nucleus accumbens and the ACC, striatum, and cerebellum^39^, and between the dorsal striatum and the middle frontal gyrus^36^. Of note, as is the case with our BOLD contrast findings, the methodology of each of those investigations was different from the present investigation in terms of population (treatment vs. non-treatment seeking), and in terms of analysis technique (ROI vs whole-brain). Interestingly, during the task, connectivity increased between the vmPFC and both the visual cortex and amygdala, when participants viewed cannabis-images, which provides early evidence that not only is there greater activation of both the visual and incentive salience circuitry, but also more connectivity between these networks. This greater connectivity is consistent with the attention bias found in substance use disorders^57^.

We found no sex differences on any of our imaging or task-based functional connectivity contrasts. Cue-reactivity studies examining sex differences in BOLD activation are limited as most studies are underpowered or do not report sex effects^58^. To our knowledge only one other study has examined sex differences in cue-induced neural activation in cannabis users^59^. In a backwards-primed subliminal cue-reactivity study Wetherill and colleagues found no sex differences in cue-induced neural activation in response to cannabis versus neutral images. However, they did demonstrate correlations between baseline craving and the bilateral insula and left lateral OFC in women, and between craving and the striatum in men. Other measures of brain function (e.g., electroencephalogram, glucose metabolism) have been used to investigate sex differences in cannabis users, and have also shown mixed results^60^. Given the notable sex differences in clinical and behavioral characteristics of cannabis use^61,62^ yet the dearth of statistically powered research on sex differences in brain function, further research in this area is warranted.

In the original validation of this fMRI task^31^, no neural activation differences were found when participants were viewing active (e.g., a picture of someone smoking cannabis products) vs. passive (e.g., a cannabis flower) images. In contrast, we found increased activation in several regions involved in complex image and facial processing (the bilateral inferior occipital cortex^63^, fusiform gyrus^64^, lateral occipital cortex^65^ and middle occipital gyrus^66^) and social cognition (left inferior parietal lobule^67^). Though these findings differed from our trial, they are likely the result of the increased sample-size of the present investigation and explained by the different content in the images (faces in the active-images and none in the passive-images).

Despite the strengths of this investigation including rigorous screening and enrollment procedures, the inclusion of a relatively large sample of participants entering treatment trials (treatment-seeking), the use of two independently recruited cohorts entering distinct treatment paradigms (potentially representing heterogenous groups of CUD participants), and, the use of data-driven whole brain analyses (to confirm regions of interest for future investigations rather than relying upon them in the present investigation), there are several limitations that warrant mention. Limitations include the fact that this was a secondary analysis of data coming from trials with slightly different procedures (for example MCQ-SF was performed prior to the scan in one trial, and following the scan for the other); and the fact that both samples were recruited from a single site in a single city with relative demographic homogeneity. These limitations may limit the generalizability of our findings, and subsequently further experimentation is needed.

In summary, we found that two separately recruited and enrolled samples of participants with CUD who were entering treatment trials displayed higher neural activation in the reward and incentive salience regions when viewing cannabis images compared to when viewing matched non-cannabis neutral images. These findings are largely consistent with the findings of other cue-reactivity trials in other SUDs, as well as those studies in cannabis users when specifically considering those with CUD (or likely CUD). We also found that participants had increased task-based connectivity between the salience network and visual and limbic systems. The largely consistent findings between these two separately recruited samples (and the consistency of this investigation with other investigations in SUDs) support the potential utility of this imaging paradigm to measure cannabis cue-reactivity in those with CUD. This paradigm may subsequently have validity as a treatment target in a similar fashion to other cue-reactivity paradigms^19^. However, further testing is needed to determine if within subject findings are consistent in terms of test-retest, and ability to change in target-engagement studies, as well as clinical trials.

## Data Availability

All data produced in the present study are available upon reasonable request to the authors

## Acknowledgements and Declaration of Interests

The authors would like to acknowledge the National Institutes of Health, Grant numbers: K23DA043628 (PI: Sahlem, NIH/NIDA), K12DA031794-04 (Co-PI’s McRae-Clark and Gray, NIH/NIDA), K24DA038240-01 (PI: McRae-Clark, NIH/NIDA), UG3DA043231 (Co-PI’s McRae-Clark and Gray, NIH/NIDA), K23 AA025399 (PI Squeglia NIH/NIAAA), K23DA045099 (PI Sherman, NIH/NIDA). We would also like to thank the many contributors to this work including Amanda Wagner, Lisa Nunn, Margaret Caruso, Taylor Rodgers, and Lauren Campbell.

## Supplemental Figures and Tables

**Supplemental-Table-1:** Significant Clusters of Activation in the combined sample. All comparisons used a voxel threshold of p<0.005, a comparison of p<0.001; a cluster significance of p<0.05, and; were family wise error (FWE) corrected. Critical T statistic for the subtraction map is 3.4491 for p <0.001, two-sided. Significant clusters had to exceed 31 voxels.

|  | Center of Mass |  |  |  |  |
| --- | --- | --- | --- | --- | --- |
| Volume | x | y | z | Mean % Signal Change | Max % Signal Change |
| Matched-neutral-images positive: |  |  |  |  |  |
| 38616 | -0.1 | -62.8 | -0.7 | 0.28 | 2.93 |
| 37 | -3.2 | 35.2 | -21.4 | 0.12 | 0.15 |
| Matched-neutral-images negative: |  |  |  |  |  |
| 2198 | 1.1 | -62.2 | 34.4 | 0.15 | -0.48 |
| 327 | 61.8 | -27.5 | 11.5 | 0.11 | -0.21 |
| 327 | -56.5 | -34.4 | 16 | 0.11 | -0.25 |
| 131 | 4.6 | 44.3 | -0.7 | 0.10 | -0.13 |
| 70 | -31.4 | -83.9 | -42.2 | 0.16 | -0.25 |
| 39 | 40.5 | -18.3 | 1 | 0.09 | -0.12 |
| 31 | -40.7 | -21.5 | 0.9 | 0.09 | -0.14 |
| Rate-positive: |  |  |  |  |  |
| 47771 | -0.5 | -48.6 | 7.4 | 0.1981 | 2.5227 |
| 76 | 28.9 | -5.1 | -33.8 | 0.1011 | 0.189 |
| 54 | 45.6 | 4.3 | -41.2 | 0.1061 | 0.1862 |
| Rate-negative: |  |  |  |  |  |
| 393 | -2.5 | 41 | -11.4 | 0.0973 | -0.1968 |
| 243 | -51.5 | -29.6 | 10.7 | 0.0728 | -0.1319 |
| 196 | -10.4 | -59.1 | 18.6 | 0.1158 | -0.2091 |
| 170 | -44.1 | -79.4 | 32.7 | 0.1452 | -0.3695 |
| 110 | 67.2 | -25.1 | 8.5 | 0.102 | -0.1891 |
| 49 | 50.1 | -72.7 | 29.2 | 0.134 | -0.2616 |
| 43 | 41.1 | -15.9 | 47.8 | 0.0757 | -0.1069 |
| 32 | -28.2 | -39.2 | -8.4 | 0.069 | -0.109 |
| Cannabis-images positive: |  |  |  |  |  |
| 40248 | -0.2 | -61.5 | -0.4 | 0.2833 | 2.7176 |
| 94 | -64.3 | -11 | -17.8 | 0.112 | 0.2758 |
| Cannabis-images negative: |  |  |  |  |  |
| 1604 | 1.4 | -65.8 | 34.4 | 0.158 | -0.4889 |
| 918 | 56.9 | -27.3 | 14.9 | 0.1314 | -0.3853 |
| 605 | -53.8 | -32.5 | 15.2 | 0.1368 | -0.3383 |
| 73 | 57.2 | -51.8 | 24.5 | 0.1222 | -0.2159 |
| 67 | -27.3 | -86.1 | -41.5 | 0.1468 | -0.2567 |
| 51 | 11.8 | -58.9 | -2.5 | 0.1523 | -0.2107 |
| 40 | -27.3 | -56.5 | 6.5 | 0.084 | -0.1403 |
| Clusters with significantly more activation during cannabis-images when compared to matched-neutral-images |  |  |  |  |  |
| 744 | -3.1 | 47.2 | -1 | 0.08 | 0.1506 |
| 559 | -1.3 | -38.9 | 28.5 | 0.0957 | 0.2368 |
| 441 | -16.5 | -98.3 | -7.3 | 0.1398 | 0.3437 |
| 299 | 18.1 | -97.4 | -2.8 | 0.1574 | 0.2805 |
| 230 | -17.9 | 44.9 | 40 | 0.0452 | 0.0762 |
| 225 | 39.7 | -44.4 | 49.1 | 0.0689 | 0.1229 |
| 205 | -44.6 | -38.9 | 50.2 | 0.0631 | 0.0912 |
| 82 | 30.6 | -68.7 | 41.3 | 0.0718 | 0.1029 |
| 68 | 19.4 | 40.3 | 49.2 | 0.0552 | 0.0917 |
| 52 | -46.1 | -64.4 | -1.8 | 0.063 | 0.0969 |
| 33 | -15.3 | 17.1 | 5.4 | 0.0448 | 0.0674 |
| Clusters with significantly <u>less</u> activation during cannabis-images when compared to matched-neutral-images |  |  |  |  |  |
| 339 | 2.4 | -87 | 27.3 | 0.0847 | -0.2491 |
| 111 | 45.8 | -30.6 | 21.7 | 0.0667 | -0.1072 |
| 36 | 13.1 | -21.3 | 41 | 0.056 | -0.0705 |

**Supplemental-Figure-1:**
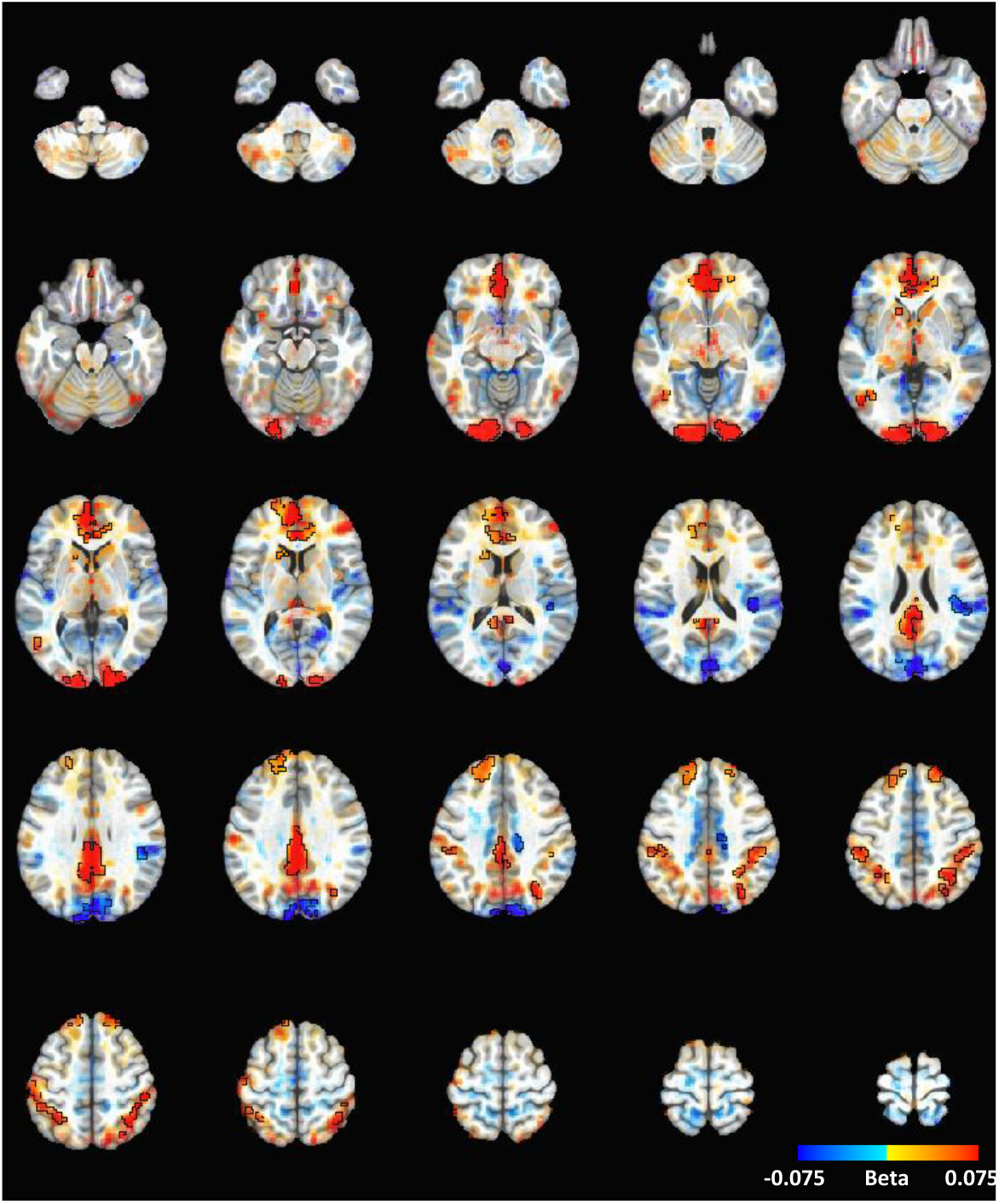
This figure displays a subtraction map of cannabis-images (both active and passive) relative to matched-neutral-images (both active and passive). All significant activation clusters are found within the black borders. Areas of activation that showed a numerical difference but did not meet statistical significance are faded based on their T-values – Group, cannabis-images vs matched-neutral-images, p <0.001, 0.05 FWE clusters.

**Supplemental-Figure-2:**
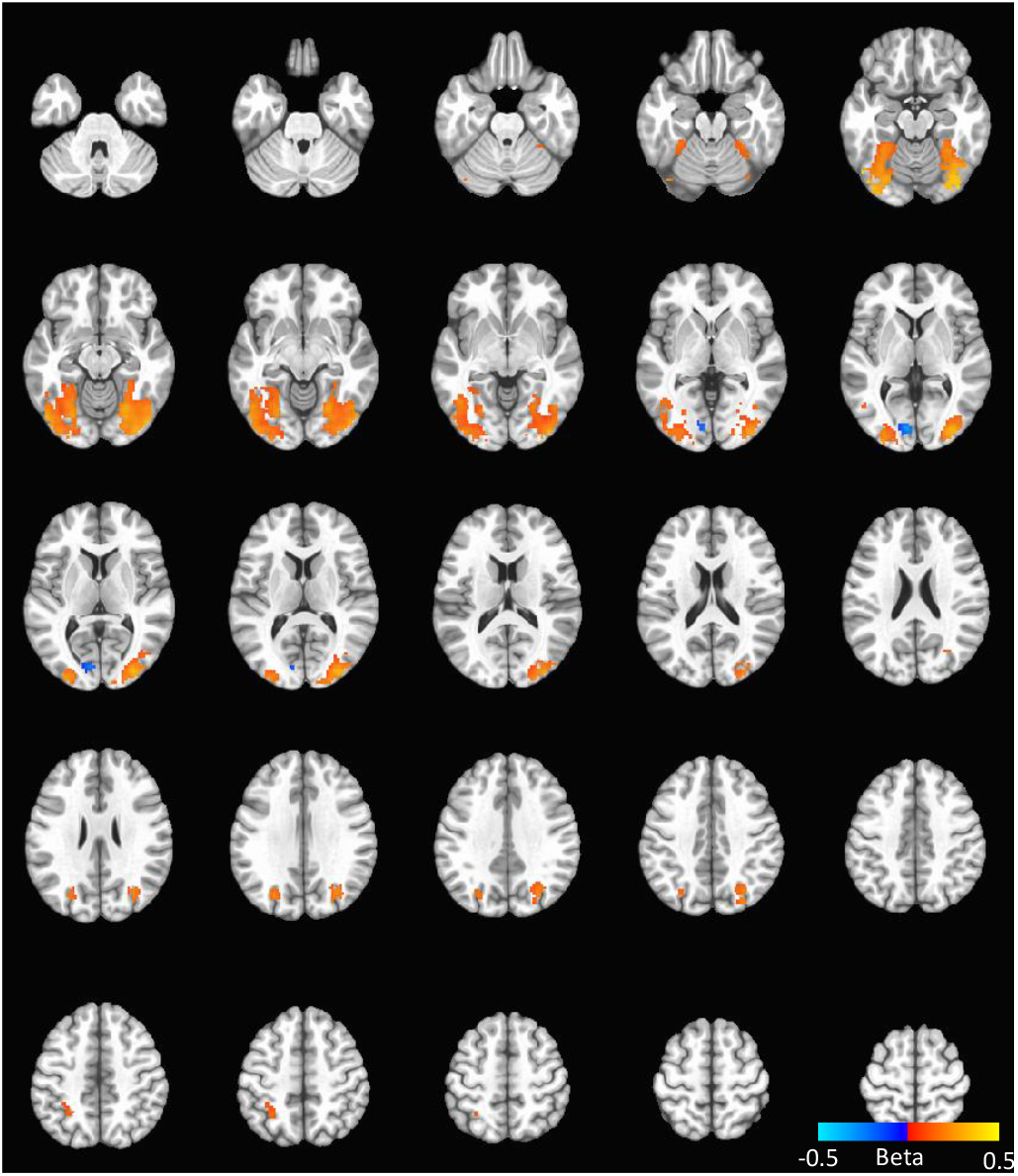
Subtraction map contrasting: (subtraction map of <u>activ</u>e cannabis images minus <u>active</u> matched neutral images) to (<u>passive</u> cannabis images minus <u>passive</u> matched neutral images [active cannabis > passive cannabis] > [active neutral > passive neutral] for the combined sample (N=65). All comparisons used a voxel threshold of p<0.001; a cluster significance of p<0.05, and; a family wise error (FWE) correction. The critical T stat is 3.4491 for p <0.001, two-sided. Clusters larger than 31 voxels are significant and shown in figure.

**Supplemental-Table-2:**
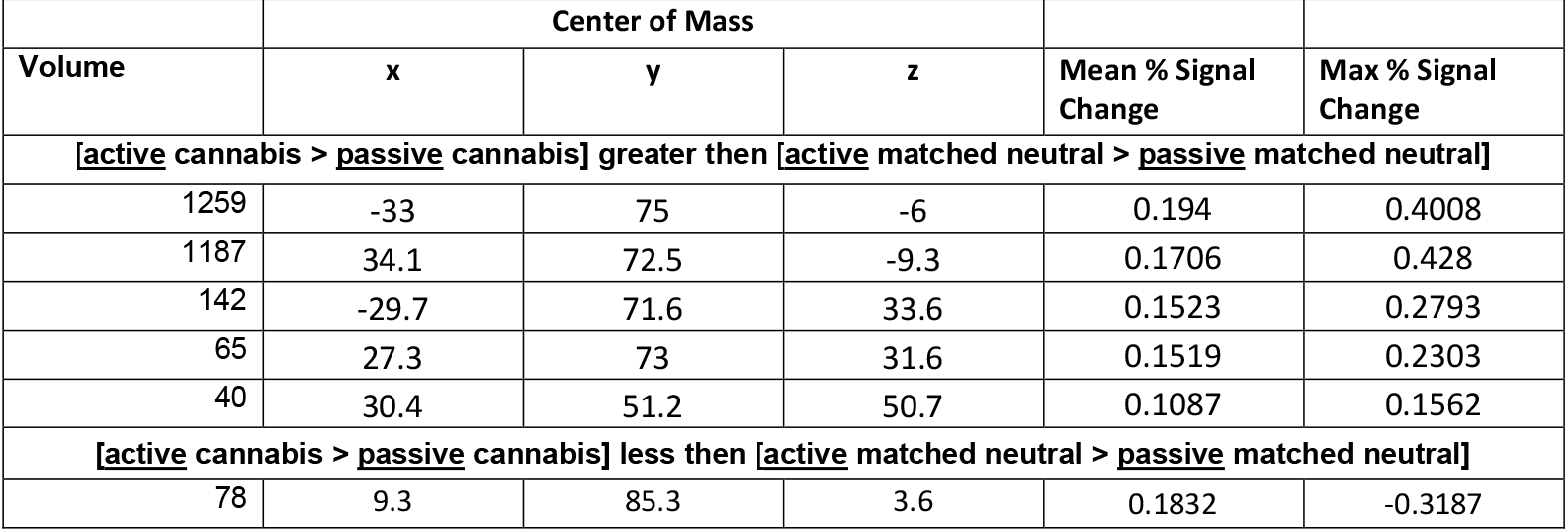
Active vs Passive images (cannabis-images vs matched-neutral-images)

**Supplemental Table 3:**
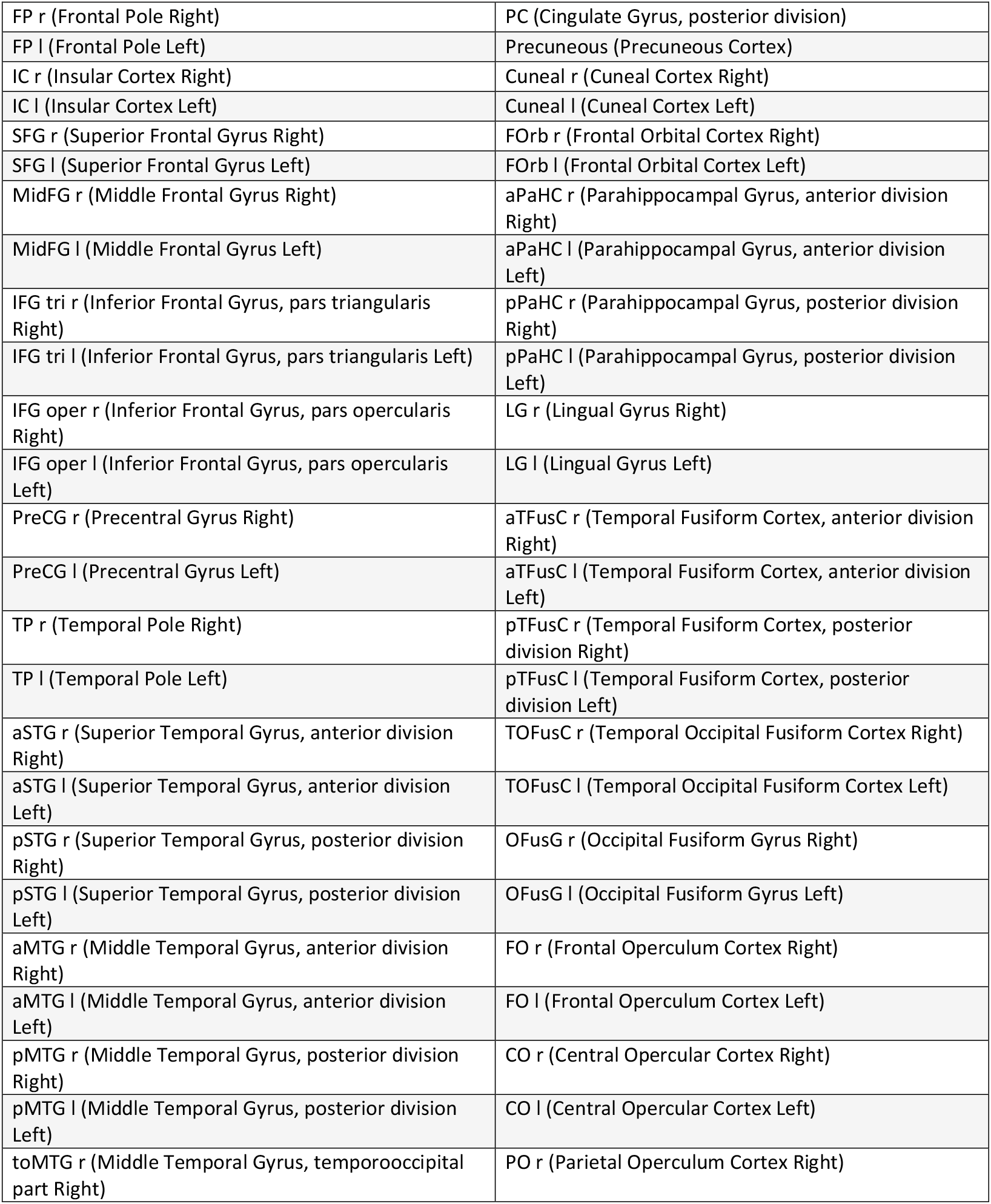
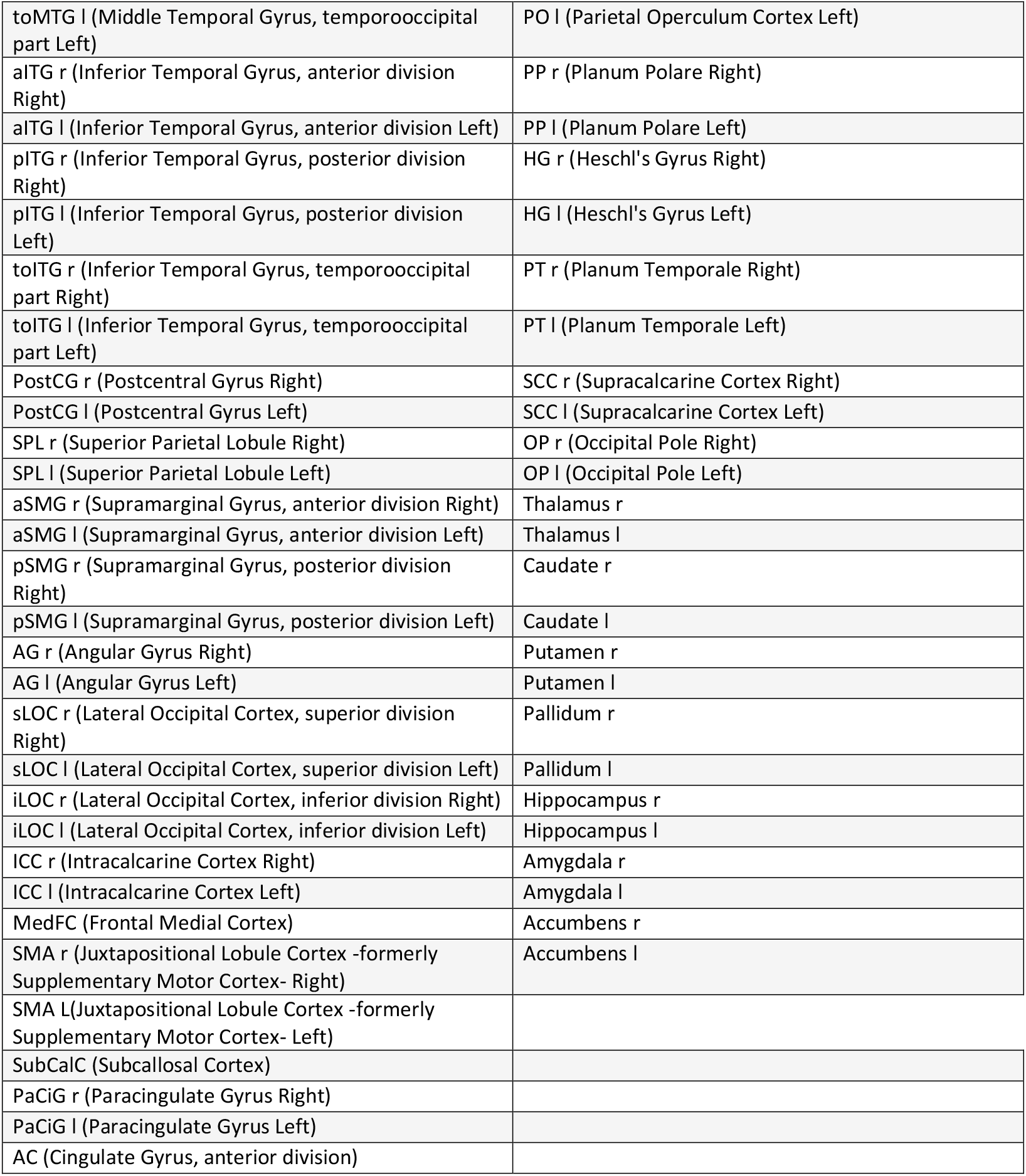
Conn Atlas regions of interest used for this study.

